# Racial and Ethnic Diversity in Clinical Studies Reported to ClinicalTrials.gov, 2009-2024

**DOI:** 10.1101/2025.07.21.25331865

**Authors:** Maryam Aziz, Emily C. O’Brien, Jay B. Lusk, Sudarshan Krishnamurthy, Vani Garcha, M. Alan Brookhart, Robert M. Califf, Michael D. Green

## Abstract

**Importance:** A lack of transparent reporting of race and ethnicity in clinical research limits the ability to identify health inequities and evaluate to what extent clinical research includes diverse populations.

**Objective:** To identify study characteristics associated with reporting race and ethnicity of clinical study participants and to document temporal trends in race and ethnicity reporting on clinicaltrials.gov.

**Design:** Cross-sectional analysis of interventional trials and observational studies from 2009-2024; multivariable logistic regression assessed study-level factors associated with reporting race and ethnicity.

**Setting:** Global registry of clinical studies (clinicaltrials.gov).

**Participants:** 58,163 studies with posted results and without early termination.

**Exposures:** Study characteristics: sponsor trial phase, study type, and country.

**Main Outcomes and Measures:** Reporting of race, reporting of ethnicity, reporting of both.

**Results:** Among 58,163 studies (mean enrollment=1,215 participants), 44.8% did not report race or ethnicity to the repository (mean enrollment=1,481 participants). The proportion of studies reporting both race and ethnicity rose from 7.4% in 2013 to 54.6% in 2024. In multivariable models, observational studies had lower odds of reporting race and ethnicity (odds ratio[OR]=0.55, 95% confidence interval[CI]=0.49–0.61) compared with interventional trials. Phase 4 trials were least likely phase to report race and ethnicity (OR=0.32; CI=0.29–0.35), and studies with only National Institute of Health funding were more likely to report race and ethnicity compared to studies with any industry funding or sponsorship (OR=1.70, CI=1.61-1.79). For studies that reported race, White participants comprised ≥50% each year based on study-level percentages; proportions of Asian participants declined, and Black participants fluctuated. ‘Not Hispanic or Latino’ remained ≥80% of reported ethnicity annually.

**Conclusions and Relevance:** Race and ethnicity reporting on clinicaltrials.gov has improved markedly yet remains incomplete, with shortfalls in late-phase and observational studies.

**KEY POINTS:** *Question:* Among clinical studies registered to clinicaltrials.gov, how often are race and ethnicity reported, how has reporting changed across time, and what study characteristics indicate a higher likelihood of reporting?

*Findings:* In a cross-sectional analysis of 58,163 studies with posted results from 2009-2024 to clinicaltrials.gov, most studies did not report race and ethnicity, though rates rose from 7% to 55%. Reporting was less common in observational and late-stage clinical trials, and more common among NIH funded studies.

*Meaning:* Despite recent gains, incomplete reporting limits generalizable assessment of clinical study participant diversity and warrants stronger oversight.

## BACKGROUND

Racial and ethnic disparities in access to care and health outcomes are well documented across a range of medical conditions.^1–3^ Despite these recognized disparities, an extensive body of literature suggests that clinical studies - the foundation of evidence-based medicine - often lack diverse participation and fail to adequately represent populations disproportionally affected by diseases.^4–6^ Many researchers and institutions have emphasized the rationale for greater diversity in clinical research.^5–8^ Without appropriate representation, study findings may lack generalizability and risk exacerbating existing disparities among underrepresented racial and ethnic groups.^9,10^ In response, several United States federal agencies launched initiatives prior to 2025 to encourage greater inclusion in clinical studies.^11,12^ Notable efforts include the National Institute of Health (NIH) Revitalization Act of 1993,^13^ and the Food and Drug Administration (FDA) 2020 guidance on Enhancing the Diversity of Clinical Trial Populations.^14^

Despite these policy efforts, comprehensive and systematic evaluations of racial and ethnic representation in clinical studies remain limited.^15^ Existing assessments tend to focus on specific diseases, time-periods, or therapeutic areas, resulting in a lack of systematic evaluation of diversity in clinical trials.^4,16–19^ Clinicaltrials.gov, the largest publicly available repository of clinical study results, includes participant characteristics such as race and ethnicity.^20^ Since 2017, the NIH Policy on the Dissemination of NIH-Funded Clinical Trial Information requires clinical studies to report race and ethnicity information to clinicaltrials.gov.^4^

To address existing gaps, we analyzed more than 58,000 interventional and observational clinical studies registered on clinicaltrials.gov from 2000 to 2024. Our objectives were two-fold:

1. To assess the state of race and ethnicity reporting to clinicaltrials.gov and examine key study characteristics associated with reporting race and ethnicity.
2. To present the distribution of participant racial and ethnic identities across key study characteristics.

## METHODS

### Data Extraction

We queried the clinicaltrials.gov registry using version 2 of the application programming interface (API) from the registry’s inception in February 2000 through December 19, 2024. This period was selected to align with the establishment of the NIH Office of Management and Budget (OMB) Race [and] Ethnicity Standard in 1997, ensuring that studies in the U.S. reported since the inception of clinicaltrials.gov in 2000 would be consistent with reporting guidelines.

We only included studies with results using the *Study Results* binary indicator retrieved using the API version 2. Upon filtering, we retrieved 58,163 studies. We obtained data for the following columns for each of the studies: *NCT ID* (the unique study identifier), *Overall Status* (status of study recruitment), *Start Date* (date first participant enrolled), *Sponsor* (the organization or person who initiates the study and has authority and control over the study), *Funder Type* (category of organization that provides funding for the study), *Primary Completion Date, Last Update Post Date, Study Type* (interventional or observational), *Phases, Enrollment, Characteristics (*demographic information collected at beginning of study). *We selected* this subset of columns because we hypothesized they may impact race and ethnicity reporting practices.

We restricted our analysis to exclude terminated trials, which are “stud[ies] [which] halted prematurely and will not resume; participants are no longer being examined or receiving intervention.”^21^ Under the Public Health Service Act, only trials with approved or licensed products that reach their primary completion date are subject to reporting results to clinicaltrials.gov. Terminated trials that are stopped before their primary completion date may have incomplete or inconsistently reported demographic results (including race and ethnicity), which could introduce misclassification bias into our analysis.^22^

Additionally we used publicly-available data from the Aggregate Analysis of ClinicalTrials.gov (AACT) database to obtain information on the country where the trials were registered.^23^ We used the January 1^st^ 2025, pipe-delimited files to obtain the “country” data which has the registered facilities/sites of the individual trial. AACT files are available at: aact.ctti-clinicaltrials.org/downloads.

### Race and Ethnicity Data Processing

The clinicaltrials.gov data was retrieved from the API version 2 in a JSON format and Python (Version 3.8.16) was used to load, parse, and convert the data into a data frame. We then used Python to iteratively extract race and ethnicity data from the *Characteristics* column, which revealed significant variability in the methods used to report race/ethnicity. In contrast, the other variables we pulled were provided in a standardized format. For race and ethnicity, some data were reported using the NIH/OMB standards, while others used customized categories and table formats. This variability resulted in the need for refinement of our data extraction script to uniformly capture race and ethnicity data. Our script returned an error for 29 entries (0.04% of studies) which contained empty tables or ‘NA’ placeholders—these entries had to be manually judged by a member of the study team.

Following the initial extraction process, we manually reviewed a random sample of 1000 rows for accuracy and completeness. The manual review uncovered 9 (0.9% of the 1000 checked) errors due to nested tables, inconsistences in table naming, incorrect data types, and studies with more than 2 race/ethnicity tables. Examples of these cases are provided as a Supplemental File. We corrected 7 of these errors by updating the logic in our Python script; however, we were unable to implement robust logic to address studies with greater than two race and ethnicity tables and with inconsistencies in table naming due to wide variation in the data structures. As a result, 115 studies (0.20% of all studies) with more than two race/ethnicity tables and 27 studies (0.05% of all studies) with inconsistencies in table naming had to be manually adjudicated by a member of the study team. Following the corrections, we performed a manual validation of 250 rows where we identified no errors.

### Race and Ethnicity Categorization

We mapped all customized race and ethnicity categories to the standardized NIH/OMB categories as defined in the Supplemental Table 1 (accessed in 2024).^24^ When the race or ethnicity was listed as “unknown or not reported”, those participants were included in our participant level tabulations. However, studies that did not report any race or ethnicity data were excluded from analyses focused on participant diversity. We used keyword search to make initial categorizations. A member of the study team (V.G.) reviewed all categorizations and manually judged any records that did not align with the predefined keywords. The NIH/OMB definitions used for categorizations are presented in Supplemental Table 1.

While all studies included in the ethnicity analysis reported some form of ethnicity data, many tables also included an “Unknown or Not Reported” category, which reflects the portion of participants for whom ethnicity is not provided. Additionally, a subset of studies (n = 1560, 2.7% of all studies) reported race and ethnicity in a single combined table. In these cases, it was unclear whether “Unknown or Not Reported” referred to race, ethnicity, or both. To avoid potential misclassification, these ambiguous entries were excluded from analyses.

### Determining Funding Source for Studies

We derived the funding source of a study based on the listed sponsors and collaborators using an approach documented elsewhere.^25–27^ The schema for determining the funding source of a study is presented in Supplemental Table 2.

### Country Classification

Using the “*countries*” pipe delimited file from the AACT database, we matched the listed country to the NCTID number for the clinical studies included in our analysis. All countries that were listed “United States”, “United States of America”, “USA”, or “US”, were considered United States based studies, and all other were considered non-United States studies. For studies with multiple countries listed, we also had a category for “at least-one US site”.

### Analysis Plan

We evaluated factors associated with the reporting of race and/or ethnicity information on clinicaltrials.gov using multivariable logistic regression. Three binary outcomes were created, 1) reporting race, 2) reporting ethnicity, and 3) reporting both. For each outcome, we fit a generalized linear model with a binomial error distribution and logit link using the RStudio glm function (RStudio 2024.04). Covariates included in the models were funder type (Industry [reference], NIH, Other, Other US Federal), trial phase (Phase 1 [reference], Early Phase 1, Phase 1/2, Phase 2, Phase 2/3, Phase 3, Phase 4, Other or Unspecified), study type (Interventional [reference] vs Observational), and study location (no United States sites [reference], at least 1 US-based site, only US based sites). We also performed a secondary analysis limited to studies conducted only in the United States to assess race and ethnicity reporting within a single regulatory environment.

Next, we examined temporal trends in race and ethnicity reporting. We presented trends in reporting both categories together and separately, as well as changes in participant diversity over time by study characteristics. To reduce the influence of larger studies on annual trends, we first calculated the within-study percentages for each race and ethnicity category, then averaged these percentages across studies for each year.

For these trends, we used the year of the study’s *Last Update Post Date,*^28^ which reflects the most recent date on which changes to the study record (including race/ethnicity) were made. We selected this date because it best reflects the point at which results are most likely to be complete. In addition, other time fields such as study registration and start date often precede the availability of results. The study’s *Last Update Post Date* values range from 2009-2024; thus, 2009 was the first year that race and ethnicity data was reported to the repository.

## RESULTS

Our results consider a total of 58,163 clinical studies, with 72.2% having at least one site in the United States. Table 1 presents key characteristics of the included studies, separated by reporting type.

**Table 1.**
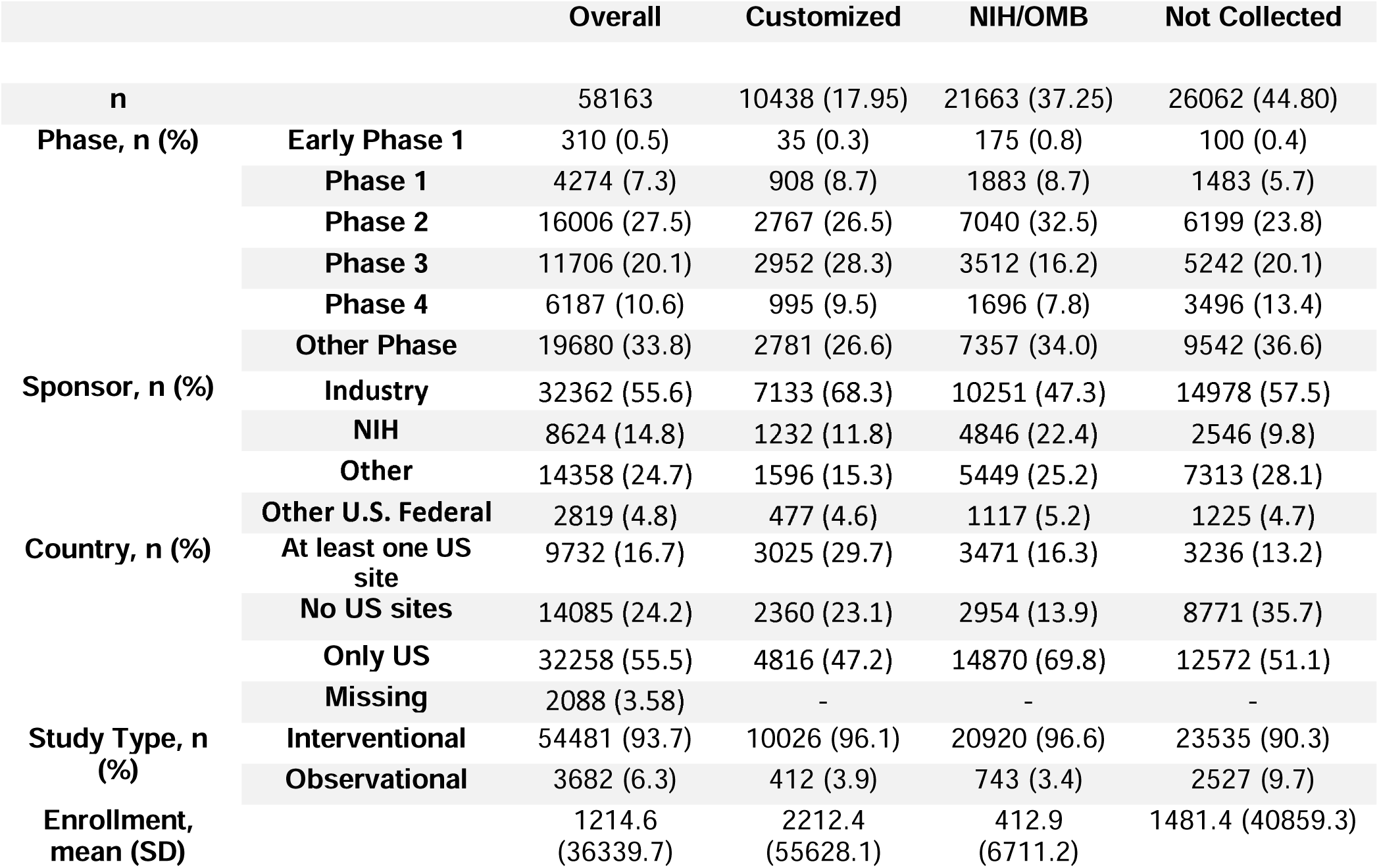
Characteristics of Clinical Trials Stratified by Race and Ethnicity Reporting Structure.

### Likelihood to Report Race and/or Ethnicity

Among the studies registered worldwide between 2009 and 2024, several design and operational characteristics were independently associated with whether investigators reported both race and ethnicity to clinicaltrials.gov (Figure 1). Compared with interventional trials, observational studies had lower odds of reporting race and ethnicity (odds ratio[OR]=0.55; 95% Confidence Interval[CI]=0.49–0.61). Phase 4 trials were the least likely of all phases to report race and ethnicity (OR=0.32,CI=0.29–0.35), and studies with other funding sources (OR=0.78, CI=0.74–0.83) were also less likely to report race and ethnicity data. In contrast, studies with only US sites (OR=5.14, CI=4.84–5.47), or with at least one site in the US (OR=5.93, CI=5.53– 6.36) were more likely to report race and ethnicity compared to those outside the US.

**Figure 1.**
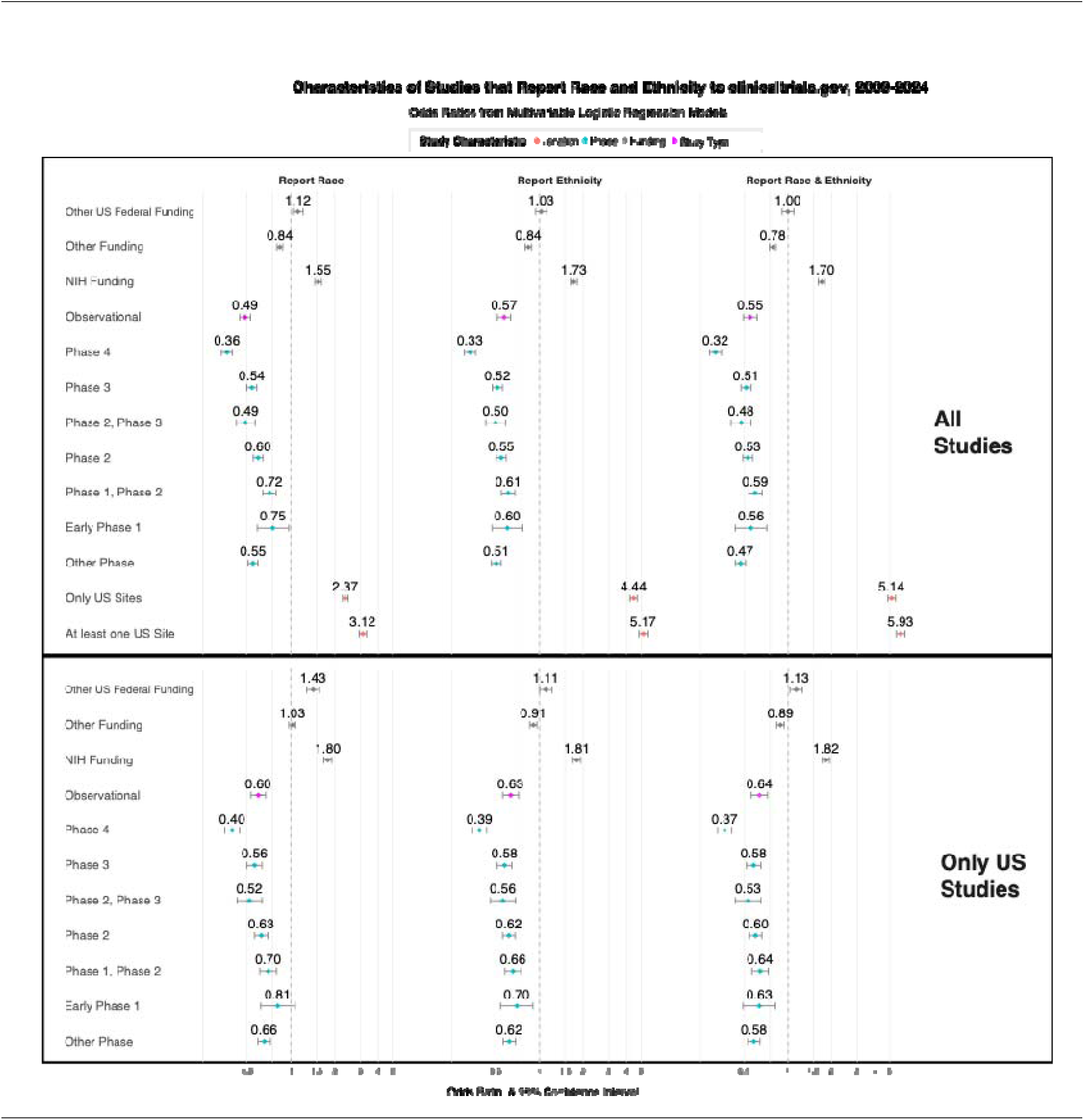
Characteristics of Studies that Report Race and Ethnicity to clinicaltrials.gov, 2009-2024. *Note*: Results produced from multivariate logistic regression. Reference category for trial phase is Phase 1. Reference category for location is non-US studies. Reference for funding is industry.

### Temporal Trends in Reporting Race and Ethnicity

Reporting of both race and ethnicity has improved over time (Figure 2). In 2024, 54.6% of all studies reported both race and ethnicity, a gradual increase from 7.4% in 2013.

**Figure 2.**
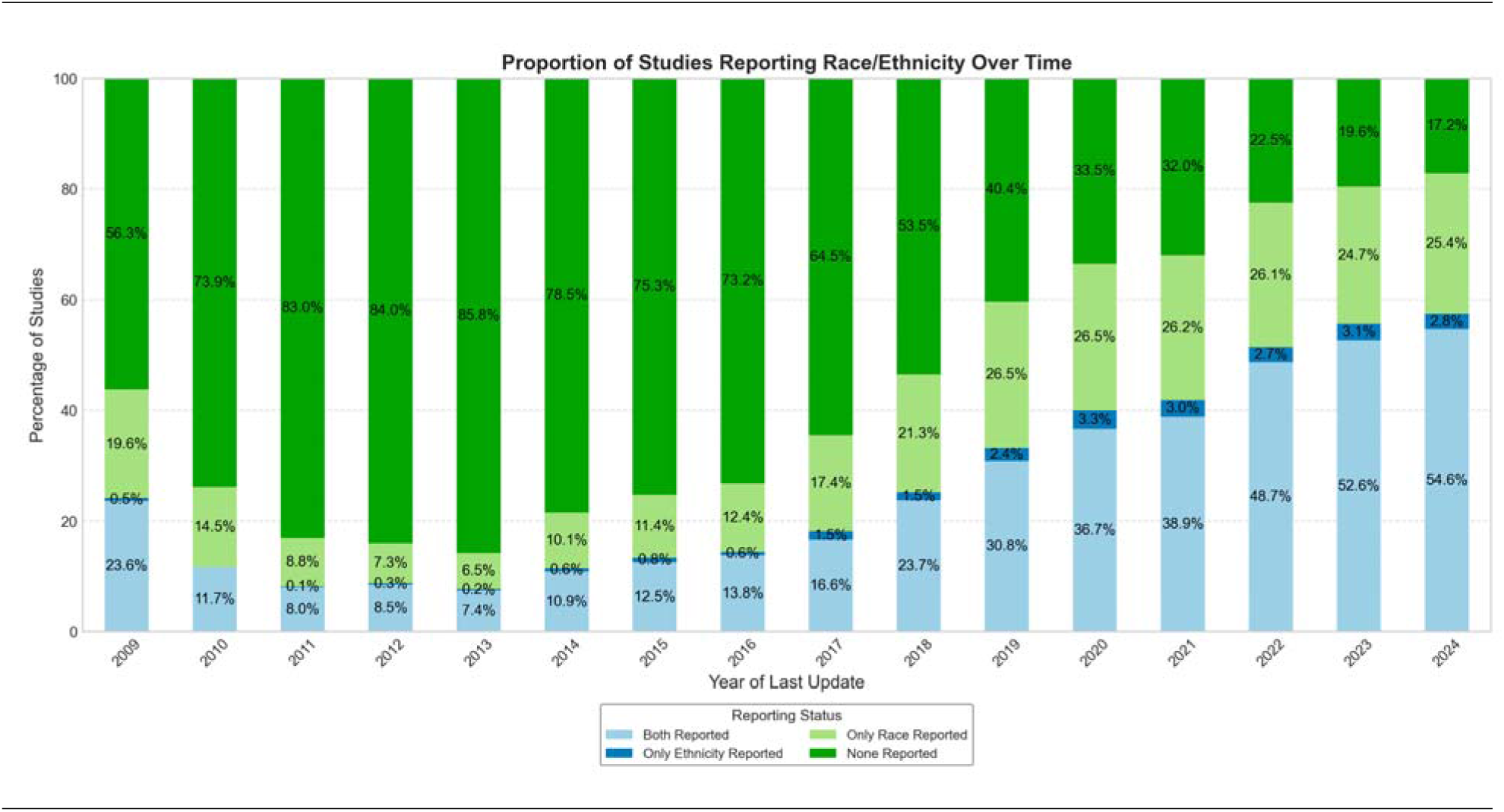
Proportion of Clinical Trials Reporting Race and Ethnicity Over Time (2009– 2024)

In studies reporting ethnicity, “Not Hispanic or Latino” was the most common category, making up 83.2% of participants in 2024 (Figure 3). While certain subgroups, including Early Phase 1 and industry-funded studies, showed higher proportions of participants identifying as “Hispanic or Latino” in certain years, these differences were not sustained (Supplemental Figures 2-12).

**Figure 3.**
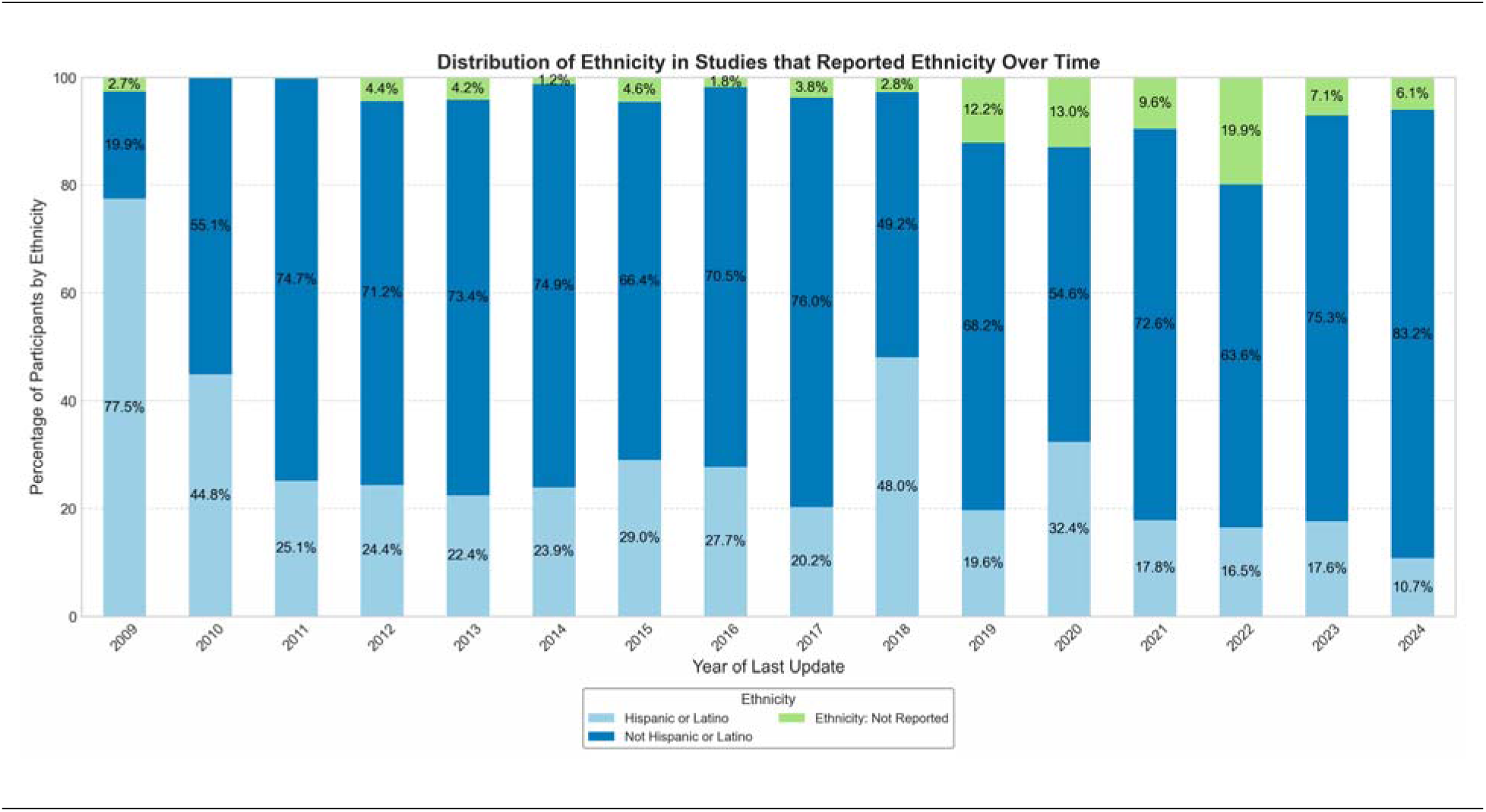
Distribution of ethnicity in studies that reported ethnicity over time (2009– 2024).

In studies that reported race, “White” was the most common category, making up more than 50% of participants each year (Figure 4). The racial distribution remained relatively stable over time, and there was a gradual decline in the proportion of “Asian” participants, while the proportion of “Black or African American” participants showed variability from year to year. When stratified by trial phase, funder type, and study design, the overall patterns persisted: “White” participants remained the majority, and changes in representation of other racial and ethnic groups were inconsistent across subgroups. Stratified results are shown in Supplemental Figures 2-12.

**Figure 4.**
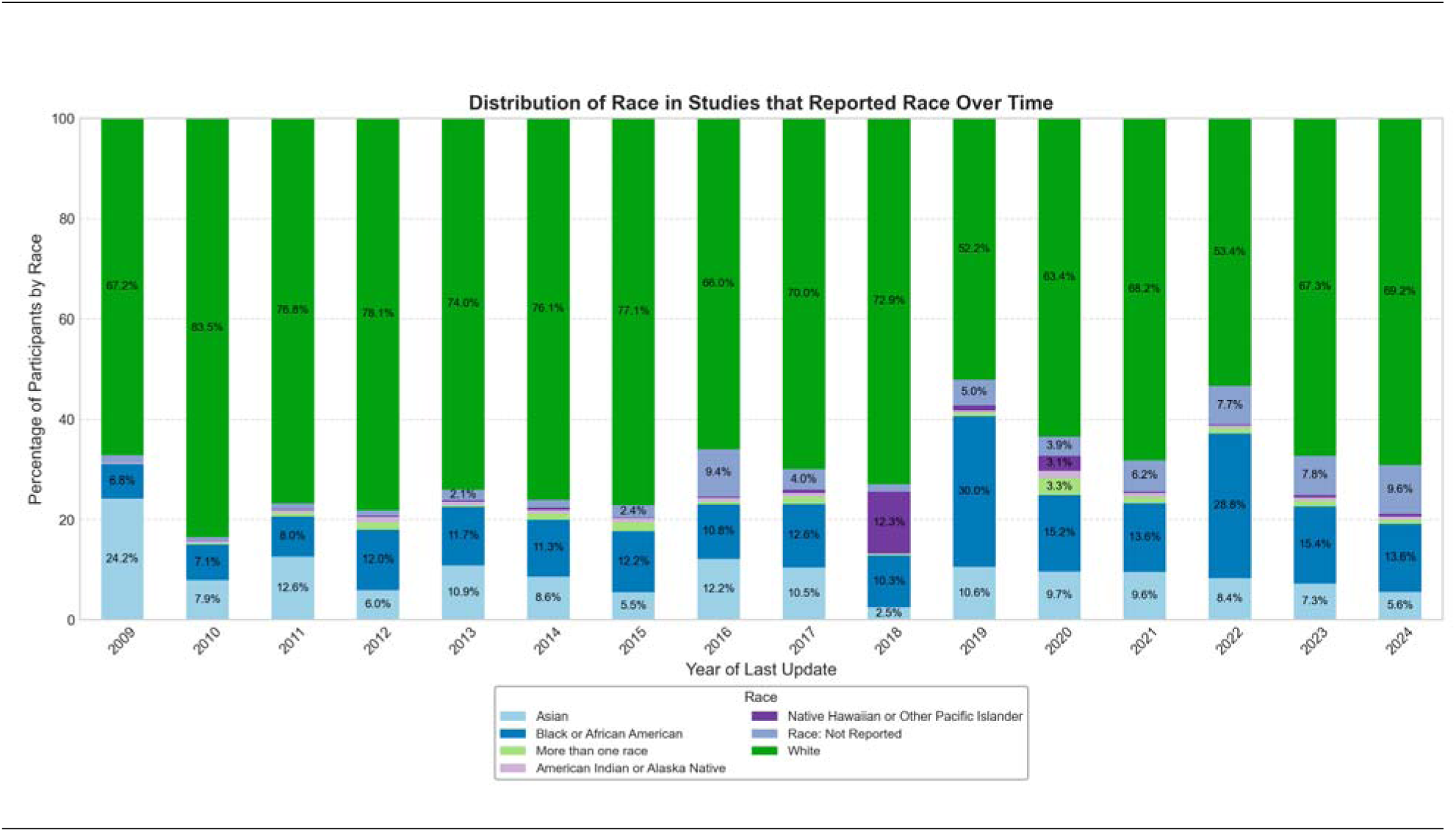
Distribution of race in studies that reported race over time (2009–2024) *Note*: The distribution of race in studies that reported race over time. For clarity, percentage labels for racial categories representing less than 2% of participants are not shown.

## DISCUSSION

To our knowledge, this is the first study to systematically document long-term trends in the reporting of race and ethnicity to clinicaltrials.gov and to quantify the characteristics of studies that are more likely to report race and ethnicity. Sufficient representation in clinical studies are essential in evaluating interventions and products across diverse populations, ensuring that they do not reinforce existing biases. In 2009, over 50% of registered studies failed to report race or ethnicity, or both; by 2024, this proportion had fallen to 17%. Early-phase trials, interventional trials, and studies that are conducted in the US were most likely to report race and ethnicity. Among US studies, the frequency of race and ethnicity reporting between federally sponsored and industry sponsored studies. However, NIH-funded studies were more likely to report both race and ethnicity.

Race is more commonly reported than ethnicity; however, in recent years over half of registered studies report both race and ethnicity, and more than three-quarters report race alone. In 2024, most adult participants were classified as either White or Black. Given that clinicaltrials.gov also includes international studies, the low and declining percentage of Asian and other racial groups is concerning, especially considering the large prevalence of Asian individuals in the US and globally. While the geographic context and racial dynamics of the countries where the research is being conducted must be considered, the availability of multilingual recruitment and reporting resources is critical to diversity initiatives.^29^ Across all study years, observational studies were consistently less likely than interventional trials to report race, ethnicity, or both. Additional work is needed to better understand why these studies are not reporting race and ethnicity data, as this information is available to study sponsors. Previous analyses have shown that trials delay reporting results to clinicaltrials.gov, despite ethical and regulatory mandates.^30^ In examples where both peer-reviewed literature and the registry are used, there are a large proportion of studies which have no available public results, despite concluding more than 7 years prior to evaluating reporting.^31^

We did not stratify by intervention type or disease area, which limits our ability to evaluate specialty-specific reporting patterns. This represents an important area for future work, as prior studies using both clinicaltrials.gov and peer-reviewed manuscripts have demonstrated variation in representation across specialties.^32,33^ Additionally, clinical trials should consider disease-specific disparities, ensuring that the composition of participants reflect the populations most affected by the condition. Clinicaltrials.gov includes more than just interventional trials, and the repository is based on the most recent updates provided by the study sponsors which can lead to some changes over time from various incentives and monitoring.^34^ The dynamic nature of the database raises the possibility of some variation in how data are recorded over time.

## Limitations

Race and ethnicity definitions differ across countries. In this study, we used NIH/OMB categories, which are standard for reporting in the United States where clinicaltrials.gov is hosted and these classifications were developed. However, we acknowledge that race and ethnicity are social constructs that depend on context, which may affect interpretation of our findings. We used a combination of keyword search, and manual review to verify that the hundreds of customized race options matched the NIH/OMB options that were presented. While both keyword searches and human review are subject to error, we are encouraged by the diminishing proportion of trials using customized methods for race and ethnicity categorization over time. Terminated trials were excluded from our study, but there is relevant concern about the representation of the individuals within these trials as well.^35^ There is increasing pharmaceutical research and development activity in low and middle income countries,^36^ which have differing racial and ethnic populations alongside interpretations of race. Our process provided an overview from our US-centric perspective. We encourage other investigators to evaluate race, ethnicity, and other systems of social hierarchy resulting in inequities in different settings. Finally, while clinicaltrials.gov is the most comprehensive public registry of trial data, it does not capture all relevant information. A previous analysis found approximately 19% of drug trials published in peer-reviewed journals were not reported to clinicaltrials.gov.^31^ There are valid concerns of missingness of race and ethnicity data and should not be considered the entire picture of clinical study representation.

## Future Directions

There are many initiatives and strategies aimed at increasing diversity in clinical trials.^6,10,37–39^ Advocacy groups, like the American Heart Association, have initiated a research network focused on the science of promoting diversity in clinical trials, with ongoing projects related to community-engagement to address barriers to clinical trial participation in minority populations, including mistrust, lack of comfort, and lack of information.^40–42^ Decentralized clinical trials have also emerged as a mechanism for providing more opportunities for underrepresented racial and ethnic populations to participate in clinical research.^43^ With the proliferation of technology and the internet, digital tools are also another promising venue for reaching more diverse populations,^44,45^ although consistent internet access remains a barrier. Collectively, these approaches for randomized controlled trials, along with other study designs,^46,47^ should be implemented in parallel to increase representation in clinical research. This includes having clear targets and priorities for improving diversity and reporting their progress towards this goal.

Since they are socially constructed categories, the meanings of race and ethnicity as well as targets for diversity in trials will change over time. We conducted this analysis to assess the current state of racial and ethnic diversity in clinical research. Transparency in reporting clinical study findings remains essential. While progress in reporting race and ethnicity data to clinicaltrials.gov may be a sign of understanding the value of this information, it is important that investigators also prioritize initiatives to consistently recruit and retain socio-demographically diverse participants.^12^ Further investigating the reporting of gender after the introduction of a new gender eligibility description registry in 2017,^48^ could provide another avenue for investigating sociodemographic diversity.

## Conclusion

We found that reporting of both race and ethnicity has improved over time, with several study characteristics associated with whether sponsors reported both participant race and ethnicity to clinicaltrials.gov. While there has been an improvement in reporting practices, more work is still needed. In 2024, 17.2% of studies still did not report any data on race and ethnicity, and 28.2% only reported either race or ethnicity. Additionally, “Not Hispanic or Latino” and “White” were the most common categories across time, with the breakdown of race and ethnicity categories not aligning with Census data across time.

Each study team has the resources, guidance, and ability to retain race and ethnicity data in some form. Emphasizing diversity in clinical trials, reporting participant sociodemographic information, and consistently implementing guidelines and policies around reporting are crucial for continued progress and improvement.

## Supporting information

Examples of custom race and ethnicity tables from clinicaltrials.gov

## Data Availability

All data are available online at clinicaltrials.gov and aact.ctti-clinicaltrials.org/downloads.

## Funding

Michael D. Green was supported by the National Institute on Aging of the National Institutes of Health under Award Number F99AG088695. The content is solely the responsibility of the authors and does not necessarily represent the official views of the National Institutes of Health.

## Competing Interests

Dr. Robert M.Califf served as the Commissioner for Food and Drug at FDA until January 2025. Prior to his appointment as FDA Commissioner, Dr Califf was an employee of and held equity in Verily Life Sciences and Google Health (Alphabet). He also served on boards of directors for Cytokinetics, Centessa, Clinetic, Keystone Symposia, the Critical Path Institute (C-Path), the Clinical Research Forum, and One Fifteen.

## Acknowledgments

We are appreciative of Annika Kruse for providing an early-stage evaluation of our idea and analysis.

## Credit Author Statement

**Maryam Aziz:** Methodology, Software, Validation, Investigation, Data Curation, Writing - Original Draft, Visualization. **Emily C. O’Brien:** Methodology, Supervision, Writing - Original Draft, Writing - Review & Editing. **Jay B. Lusk:** Writing - Review & Editing. **Sudarshan Krishnamurthy:** Writing - Review & Editing. **Vani Garcha:** Data Curation, Writing - Review & Editing. **M. Alan Brookhart:** Conceptualization, Writing - Review & Editing, Supervision. **Robert M. Califf:** Conceptualization, Writing - Review & Editing, Supervision. **Michael D. Green:** Conceptualization, Methodology, Formal analysis, Investigation, Data Curation, Writing - Original Draft, Writing – Review and Editing, Supervision, Project administration, Funding acquisition

## Notes

### Author Declarations

We queried the clinicaltrials.gov registry using version 2 of the application programming interface (API) from the registry inception in February 2000 through December 19, 2024. Additionally we used publicly-available data from the Aggregate Analysis of ClinicalTrials.gov (AACT) database to obtain information on the country where the trials were registered. We used the January 1st 2025, pipe-delimited files to obtain the country data which has the registered facilities/sites of the individual trial. AACT files are available at: aact.ctti—clinicaltrials.org/downloads.

