## Supplemental Tables and Figures for "Racial and Ethnic Diversity in Clinical Studies Reported to ClinicalTrials.gov, 2009-2024"

**Supplemental Table 1. NIH/OMB Race and Ethnicity Categories**

| Race (NIH/OMB) | Category | Definition |
| --- | --- | --- |
|  | American Indian or Alaska Native | A person having origins in any of the original peoples of North and South America (including Central America), and who maintains tribal affiliation or community attachment. |
|  | Asian | A person having origins in any of the original peoples of the Far East, Southeast Asia, or the Indian subcontinent including, for example, Cambodia, China, India, Japan, Korea, Malaysia, Pakistan, the Philippine Islands, Thailand, and Vietnam. |
|  | Black or African American | A person having origins in any of the black racial groups of Africa. Terms such as "Haitian" or "Negro" can be used in addition to "Black or African American." |
|  | Native Hawaiian or Other Pacific Islander | A person having origins in any of the original peoples of Hawaii, Guam, Samoa, or other Pacific Islands. |
|  | White | A person having origins in any of the original peoples of Europe, the Middle East, or North Africa. |
|  | More than one race | More than one race |
|  | Unknown or not reported | Unknown or not reported |
| Ethnicity (NIH/OMB) | Hispanic or Latino | A person of Cuban, Mexican, Puerto Rican, South or Central American, or other Spanish culture or origin, regardless of race. The term, "Spanish origin," can be used in addition to "Hispanic or Latino.” |
|  | Not Hispanic or Latino | Not Hispanic or Latino |
|  | Unknown or not reported | Unknown or not reported |

**Supplemental Table 2. How Funding Source was Identified**

| **Funding Source** | **Lead Sponsor** | **Collaborators** |
| --- | --- | --- |
| **Industry** | **Industry** | **Any type** |
|  | **Other** | **At least one from Industry, none from NIH/Other U.S. Fed** |
| **NIH** | **NIH** | **Any type** |
|  | **Other U.S. Fed** | **At least one from NIH** |
|  | **Other** | **At least one from NIH** |
| **Other U.S. Fed** | **Other U.S. Fed** | **None from NIH** |
|  | **Other** | **At least one from Other U.S. Fed, none from NIH** |
| **Other** | **Other** | **None from Industry, NIH, Other U.S. Fed** |

**Supplemental Figure 1. Proportion of Trials Reporting Race and Ethnicity Over Time, Stratified by Study Type**

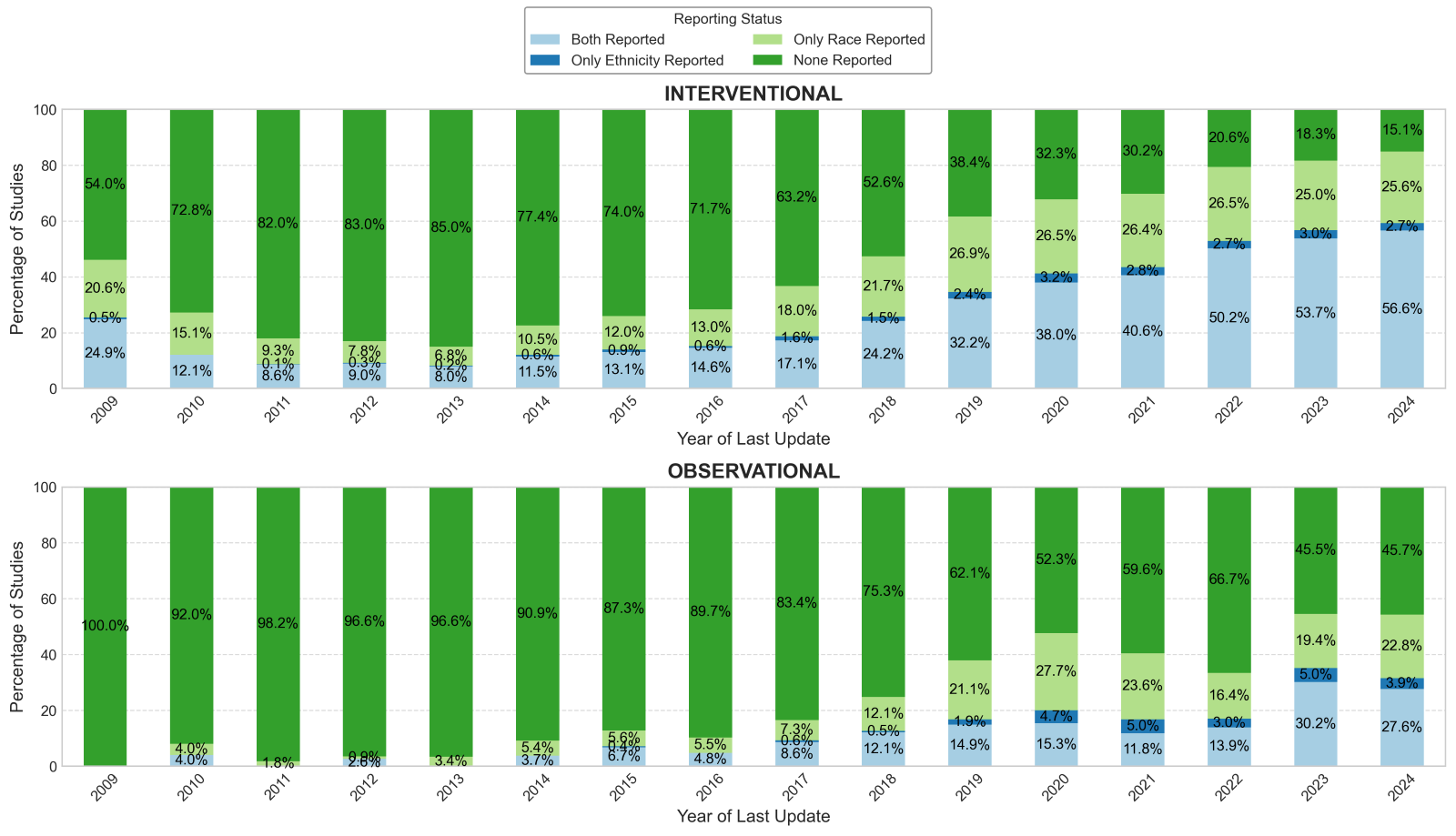

**Supplemental Figure 2. Proportion of Trials Reporting Race and Ethnicity Over Time, Stratified by Study Phase**

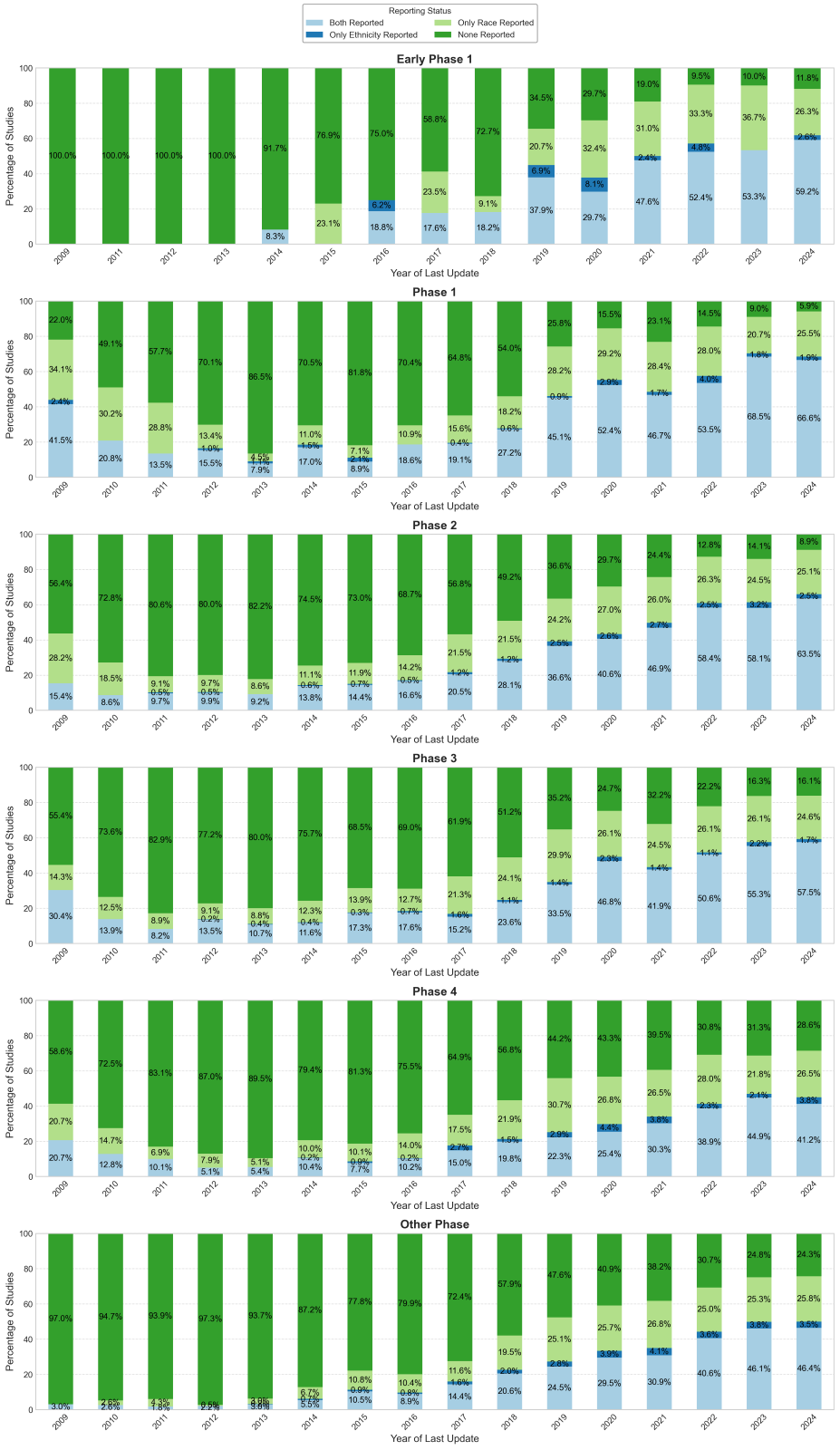

**Supplemental Figure 3. Proportion of Trials Reporting Race and Ethnicity Over Time, Stratified by Sponsor Type**

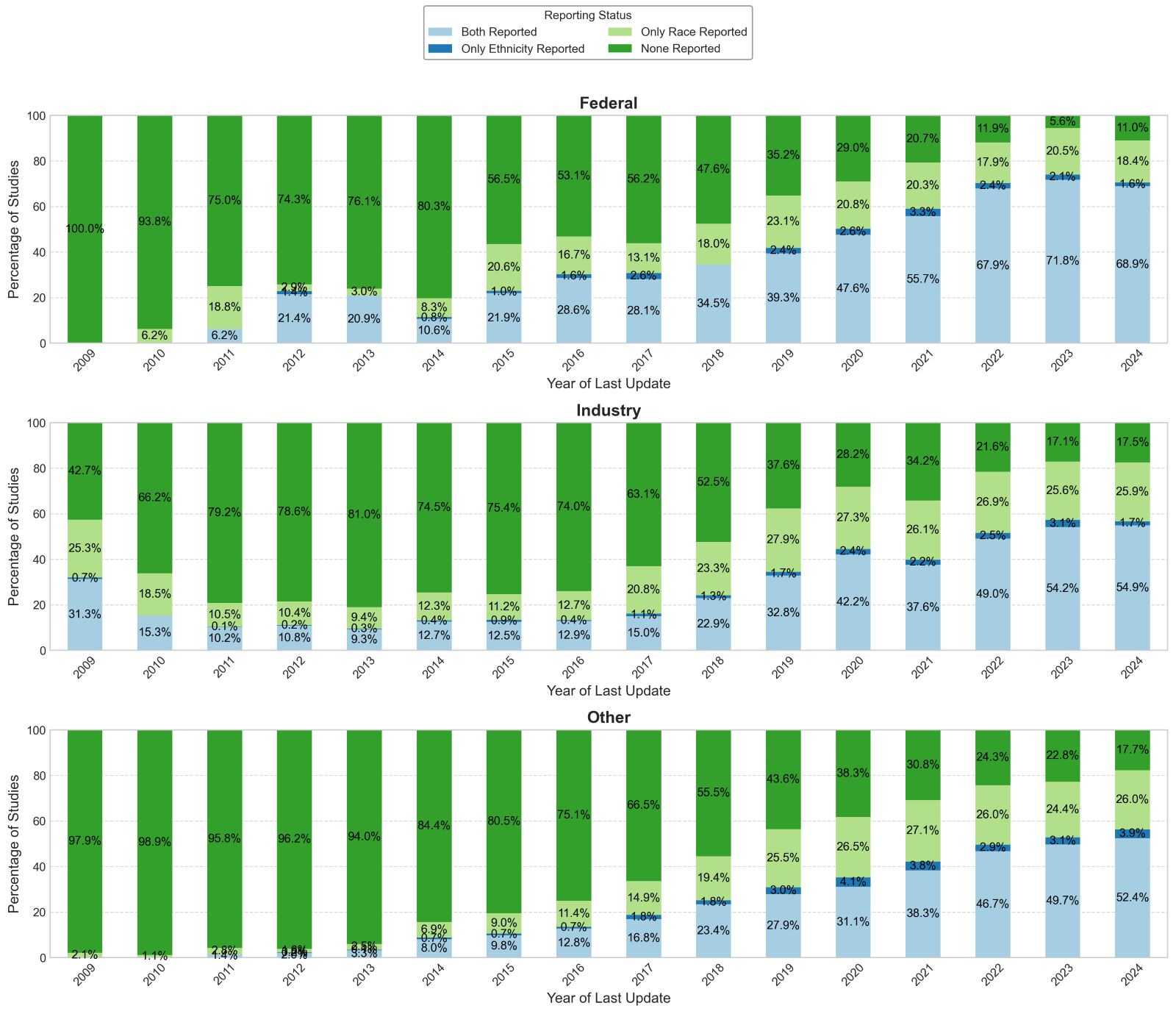

**Supplemental Figure 4. Proportion of Trials Reporting Race and Ethnicity Over Time, Stratified by US vs Non-US**

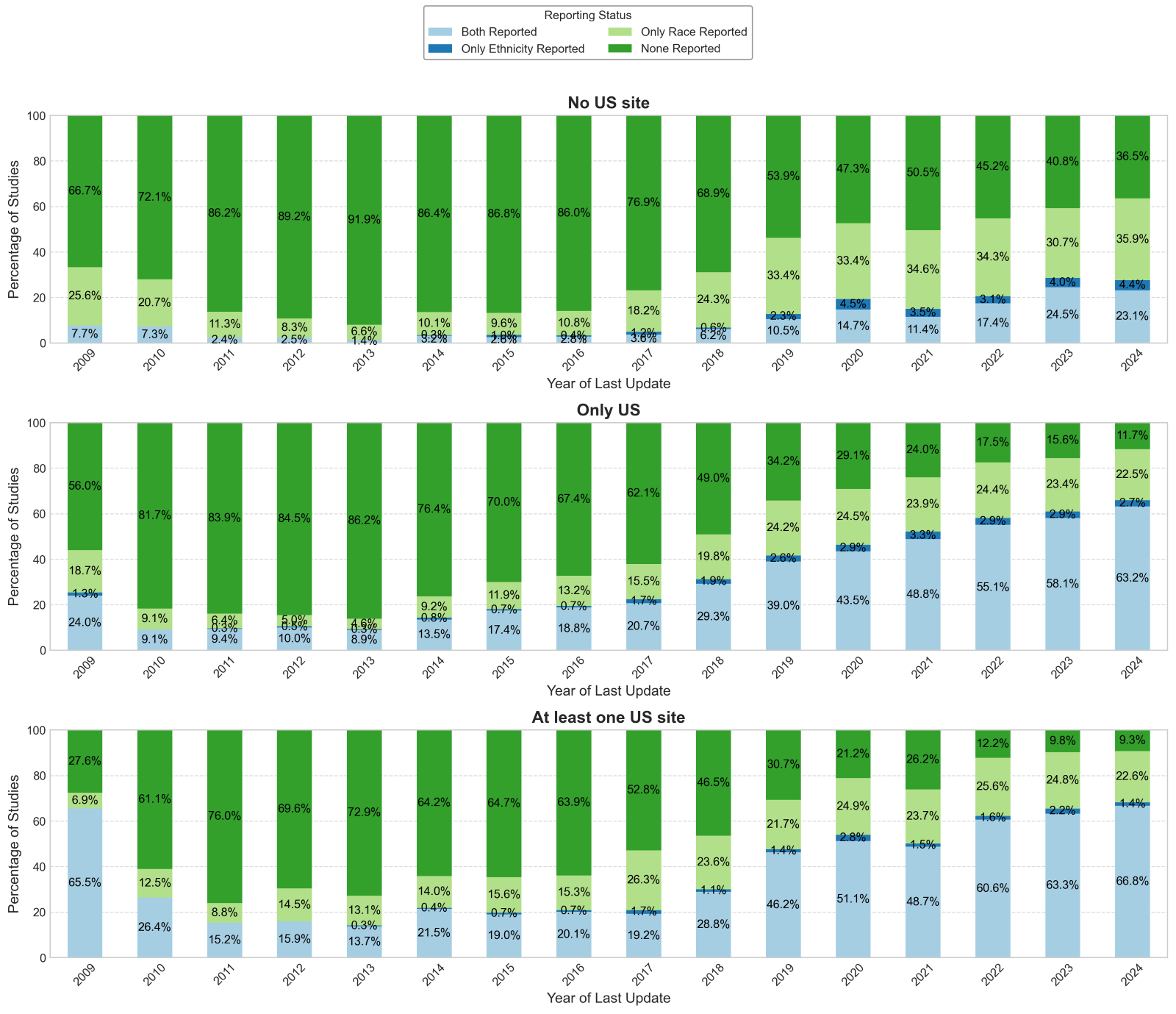

**Supplemental Figure 5. Distribution of ethnicity in studies that reported ethnicity over time, Stratified by Study Type**

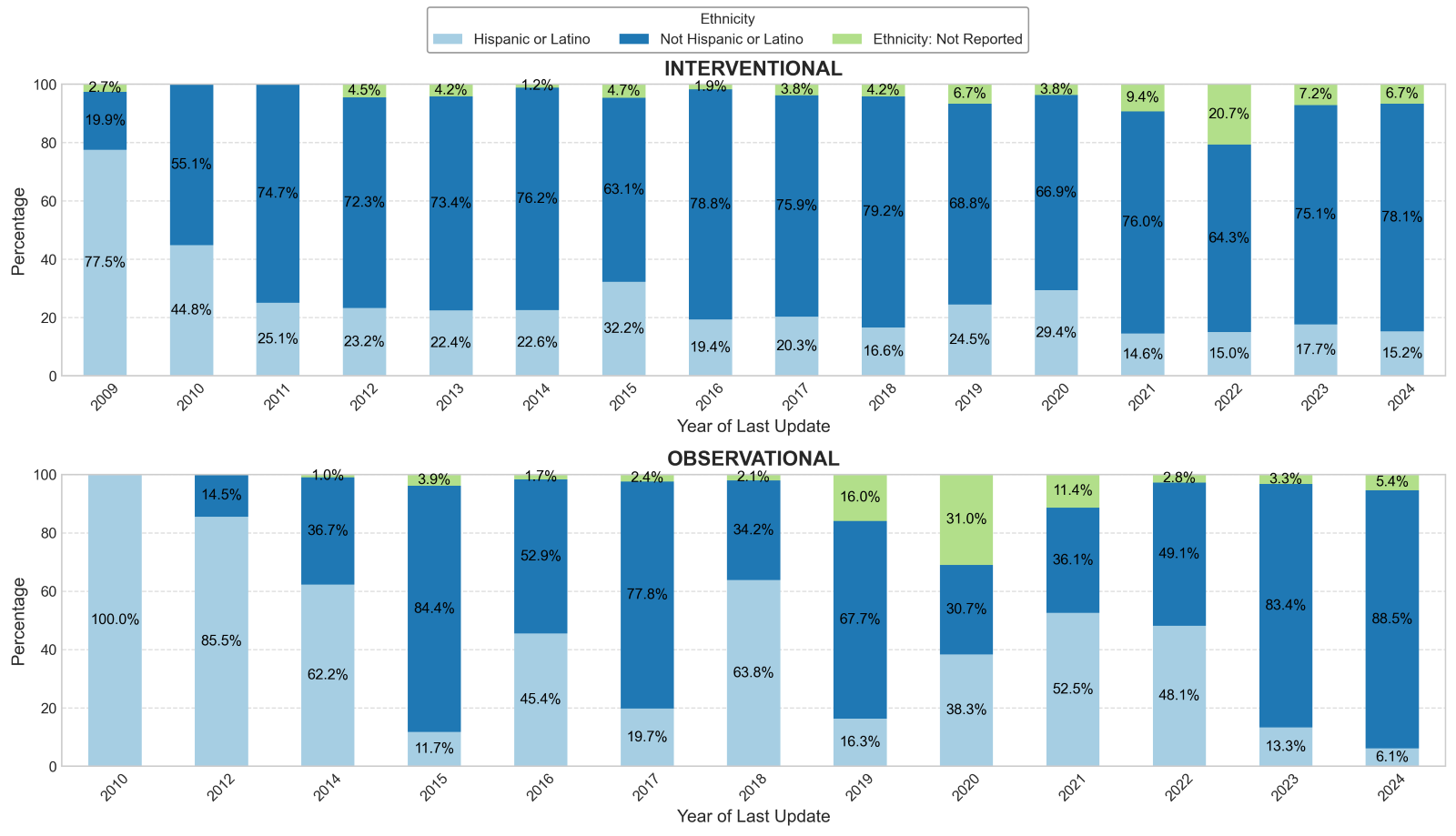

**Supplemental Figure 6. Distribution of ethnicity in studies that reported ethnicity over time, Stratified by Study Phase**

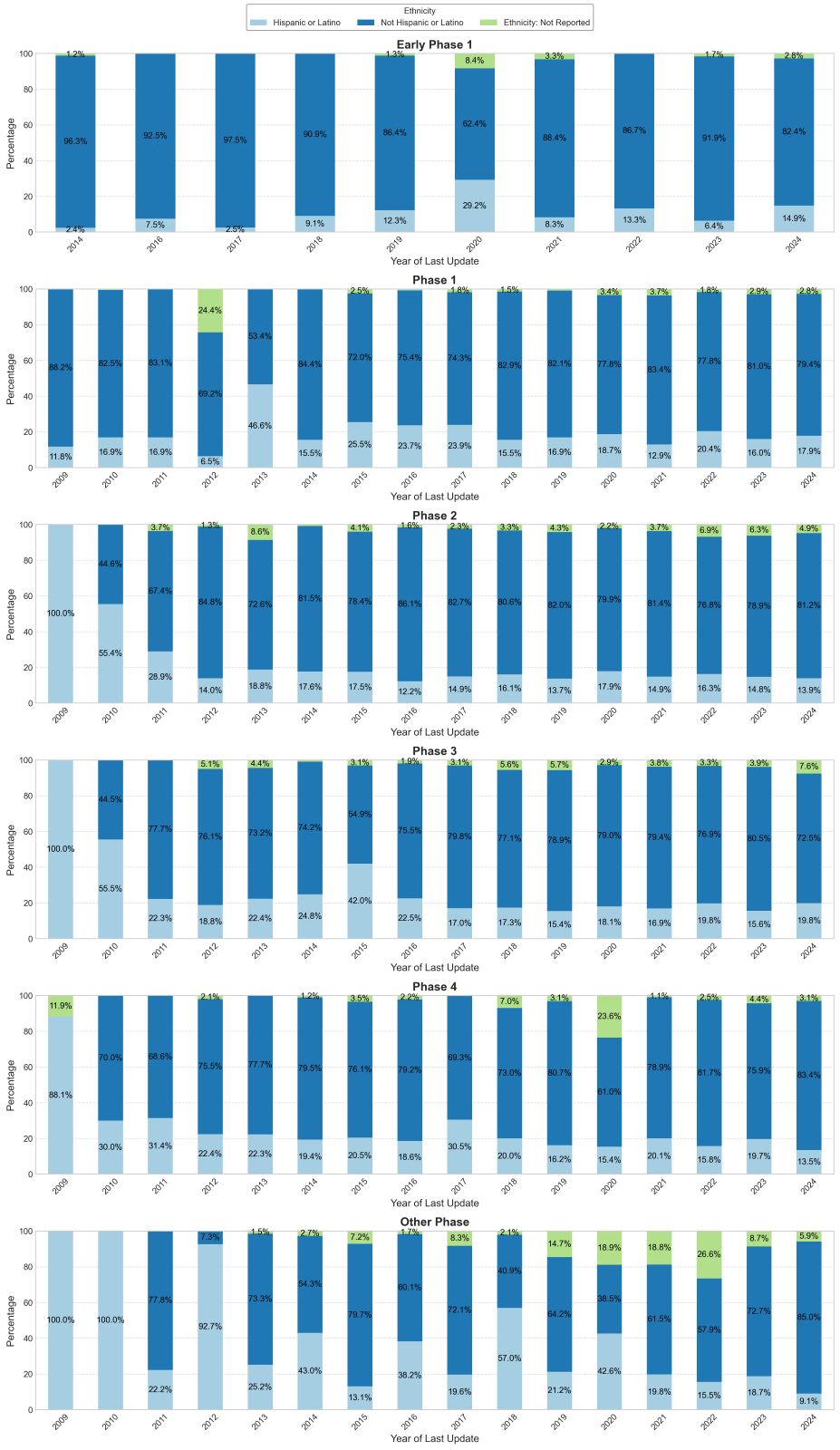

**Supplemental Figure 7. Distribution of ethnicity in studies that reported ethnicity over time, Stratified by Sponsor Type**

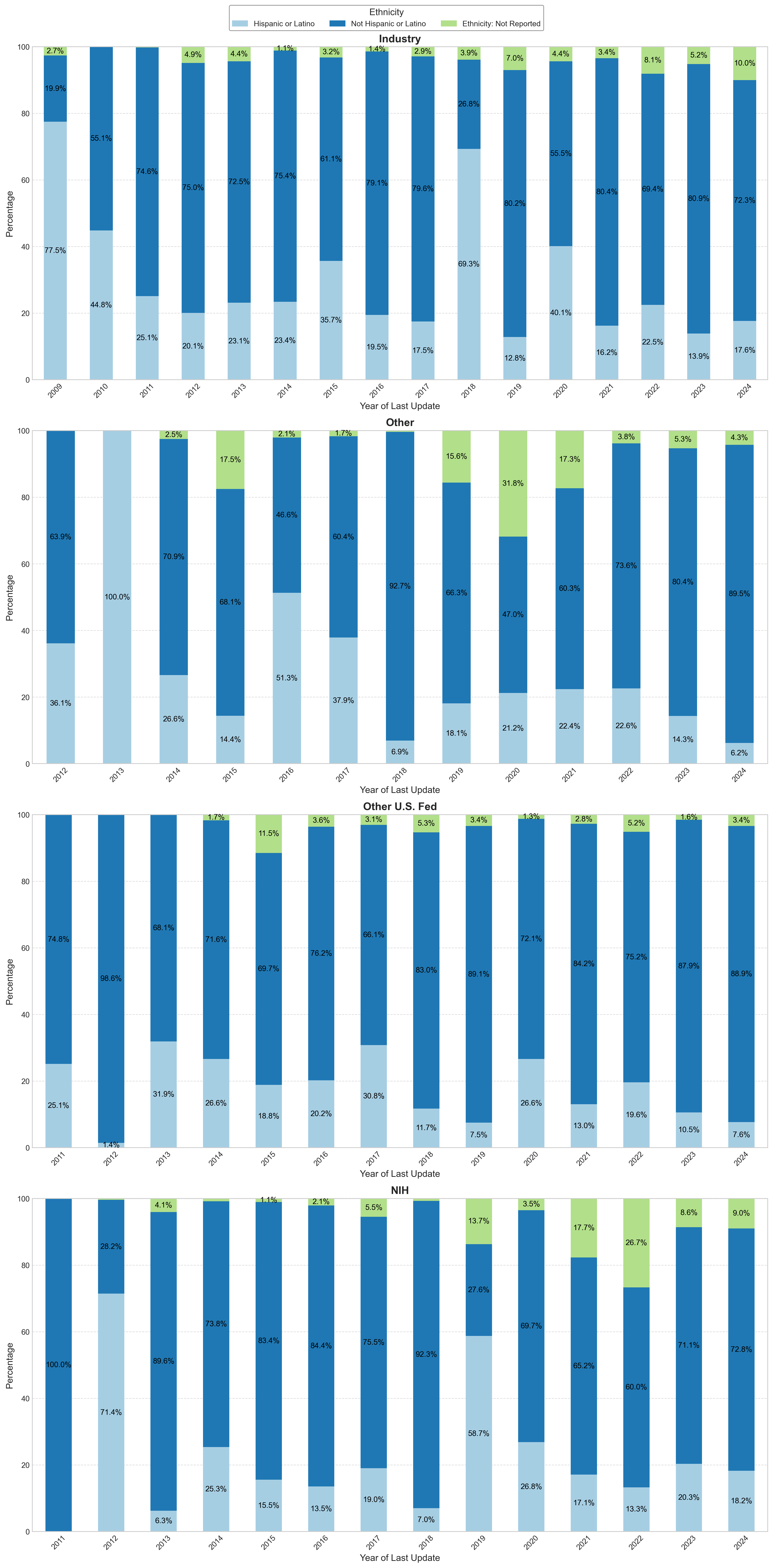

**Supplemental Figure 8. Distribution of ethnicity in studies that reported ethnicity over time, Stratified by US vs Non-US**

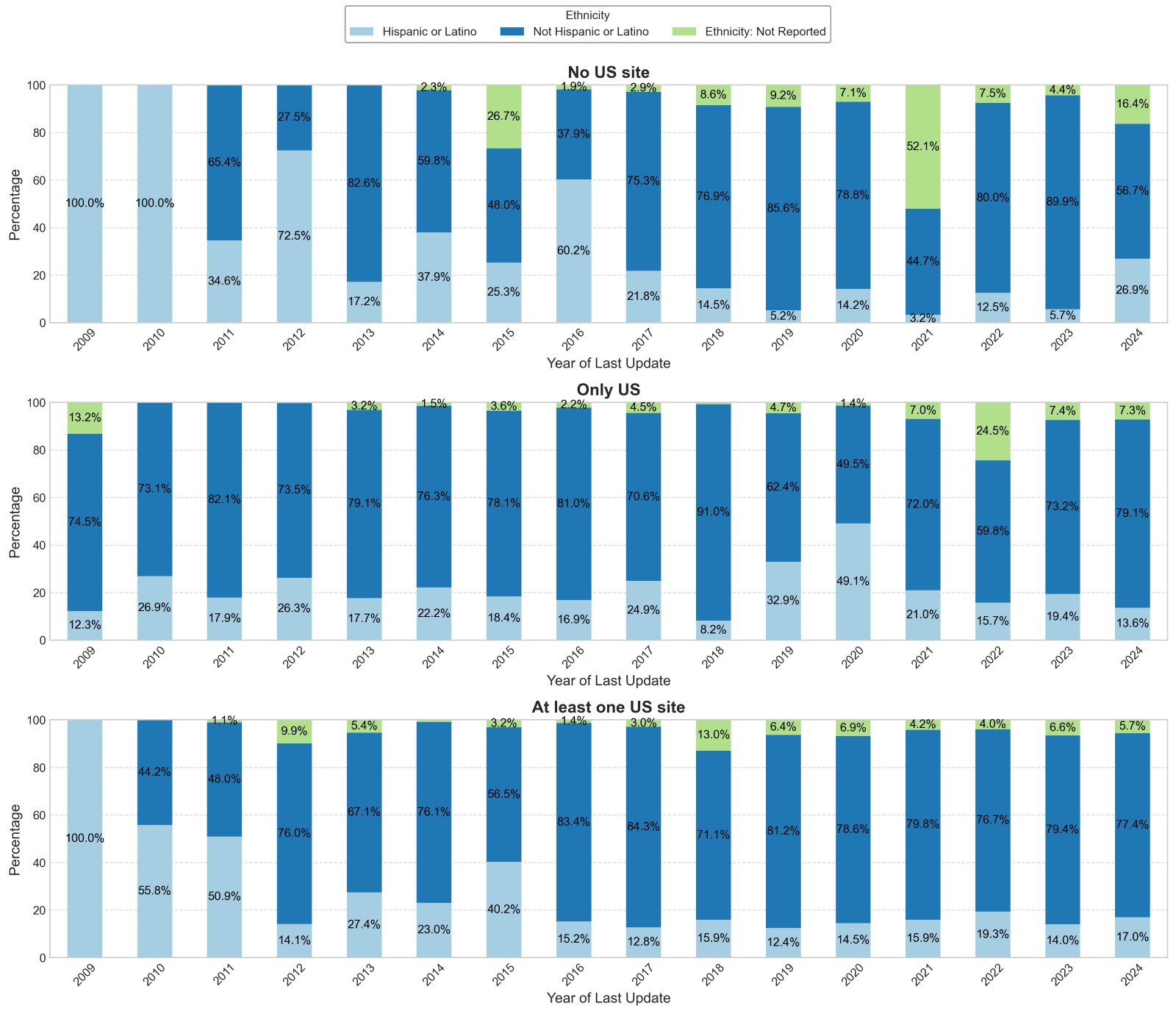

**Supplemental Figure 9. Distribution of race in studies that reported race over time, Stratified by Study Type**

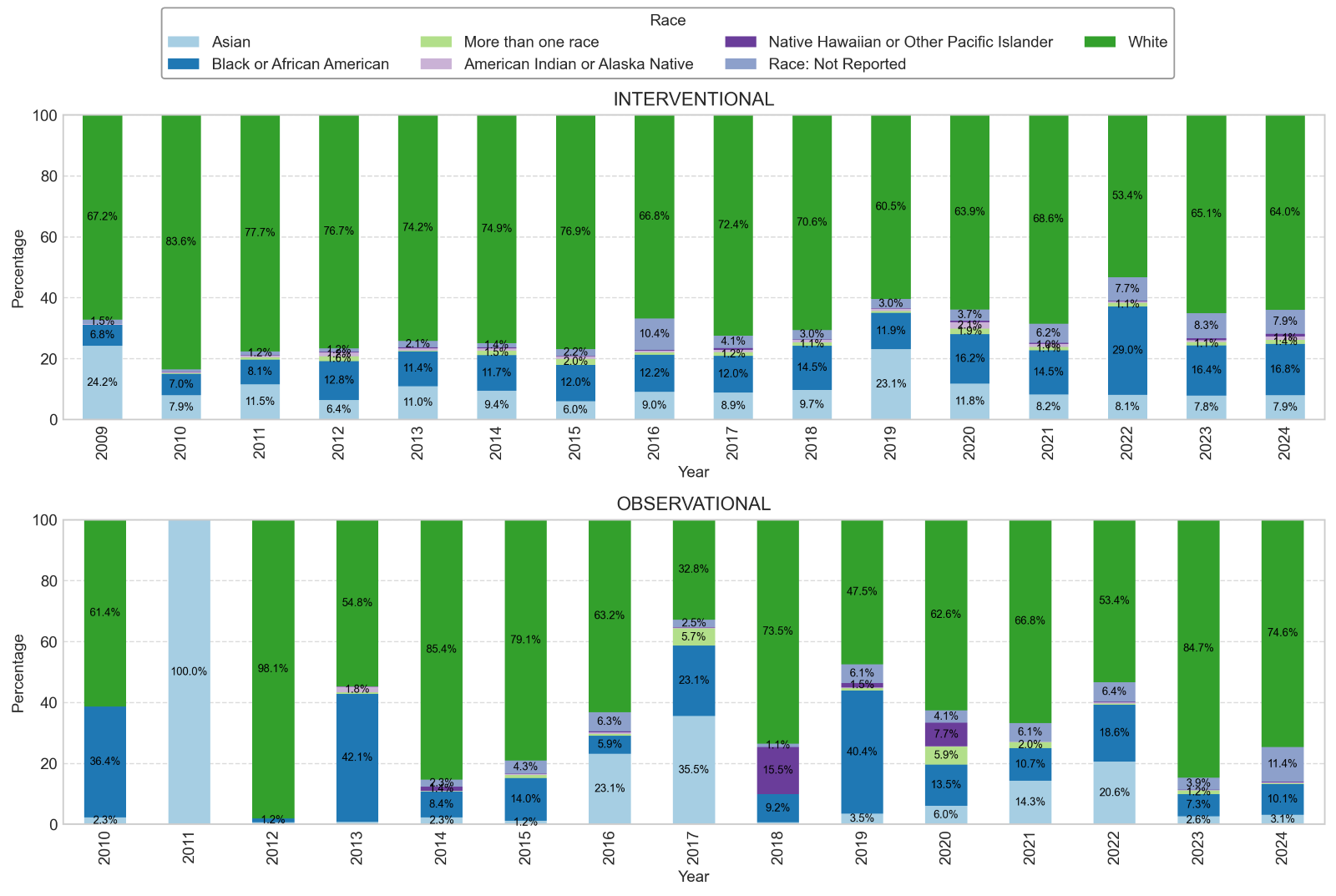

**Supplemental Figure 10. Distribution of race in studies that reported race over time, Stratified by Study Phase**

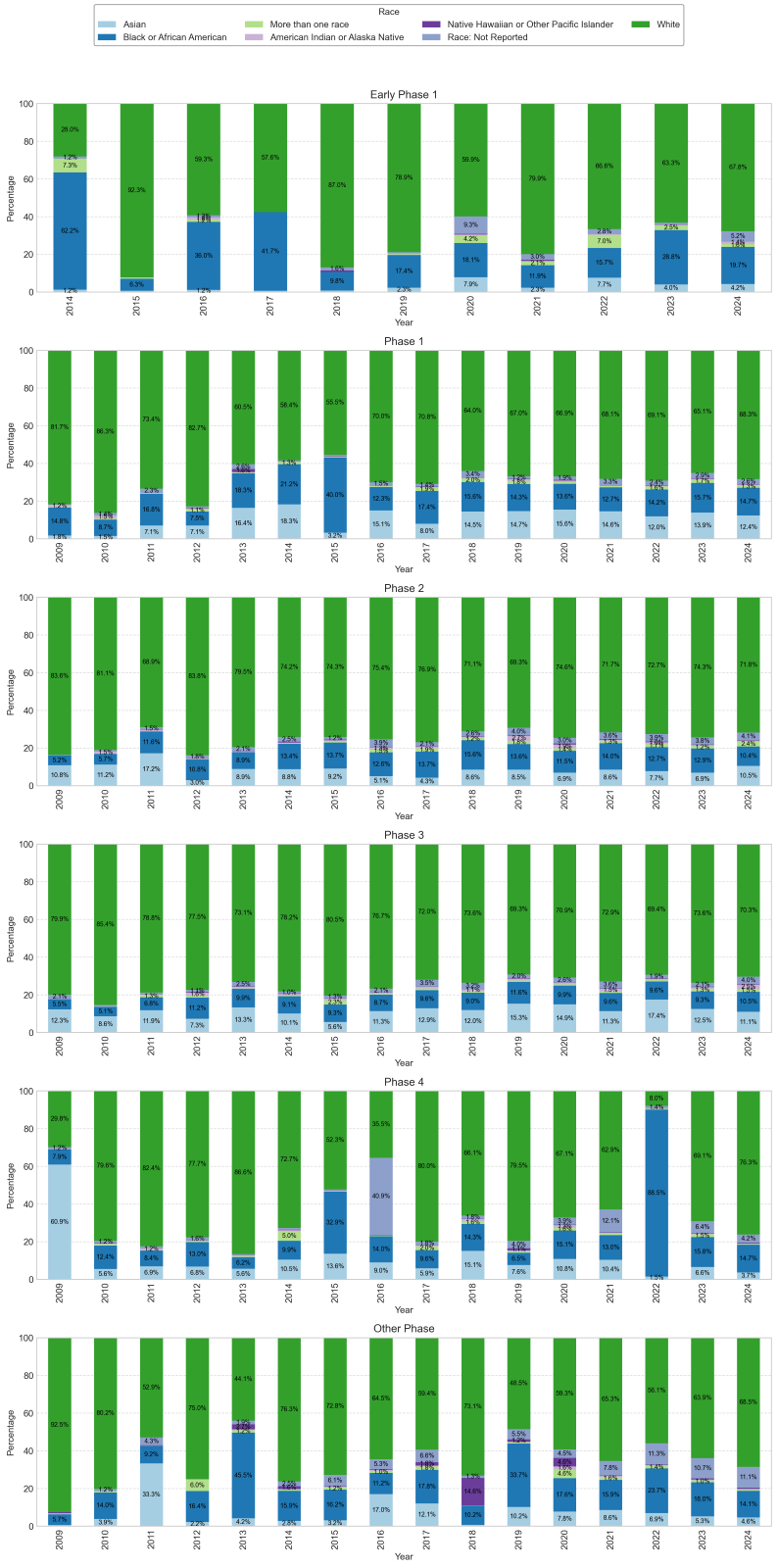

**Supplemental Figure 11. Distribution of race in studies that reported race over time, Stratified by Sponsor Type**

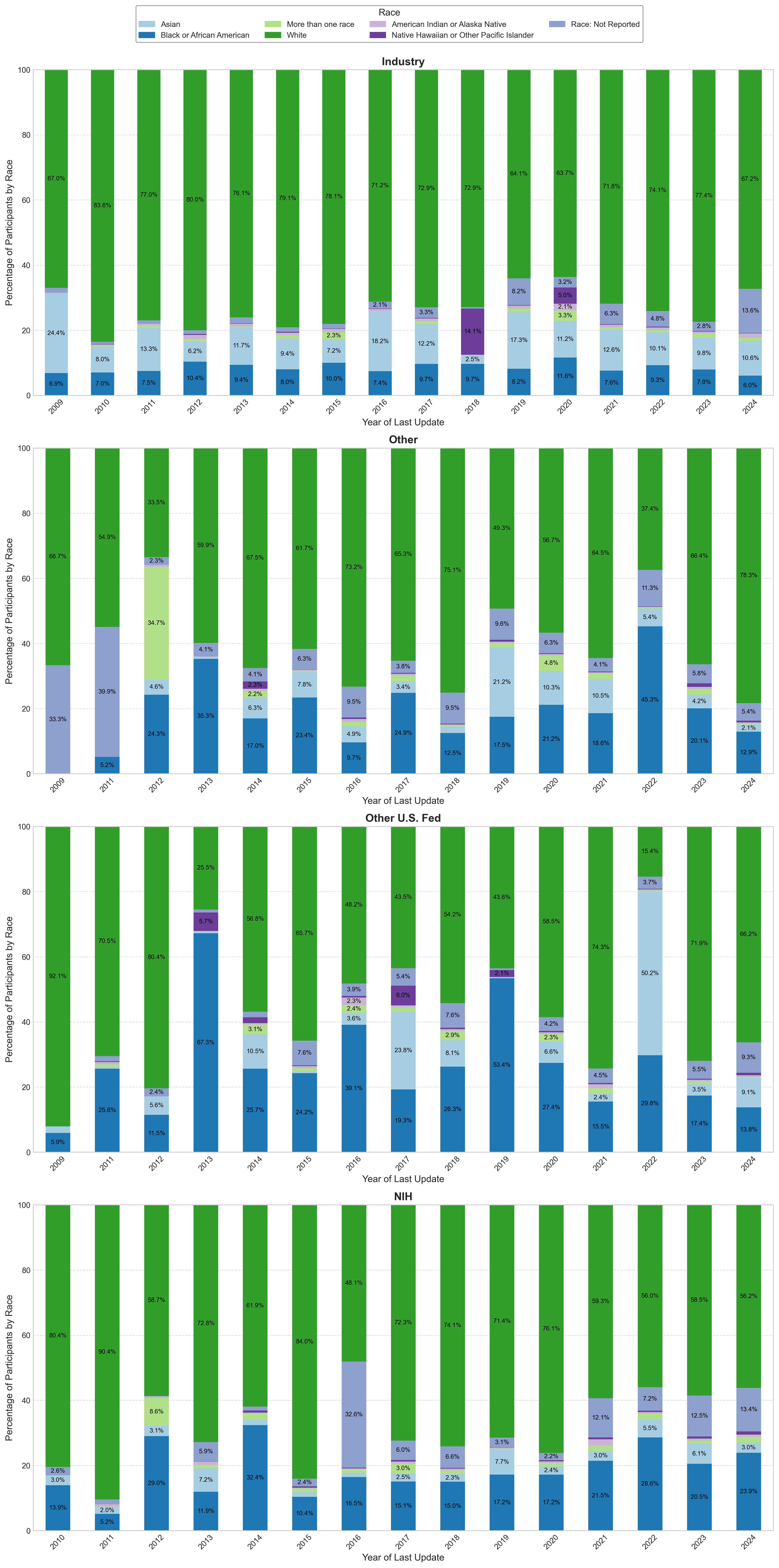

**Supplemental Figure 12. Distribution of race in studies that reported race over time, Stratified by US vs Non-US**

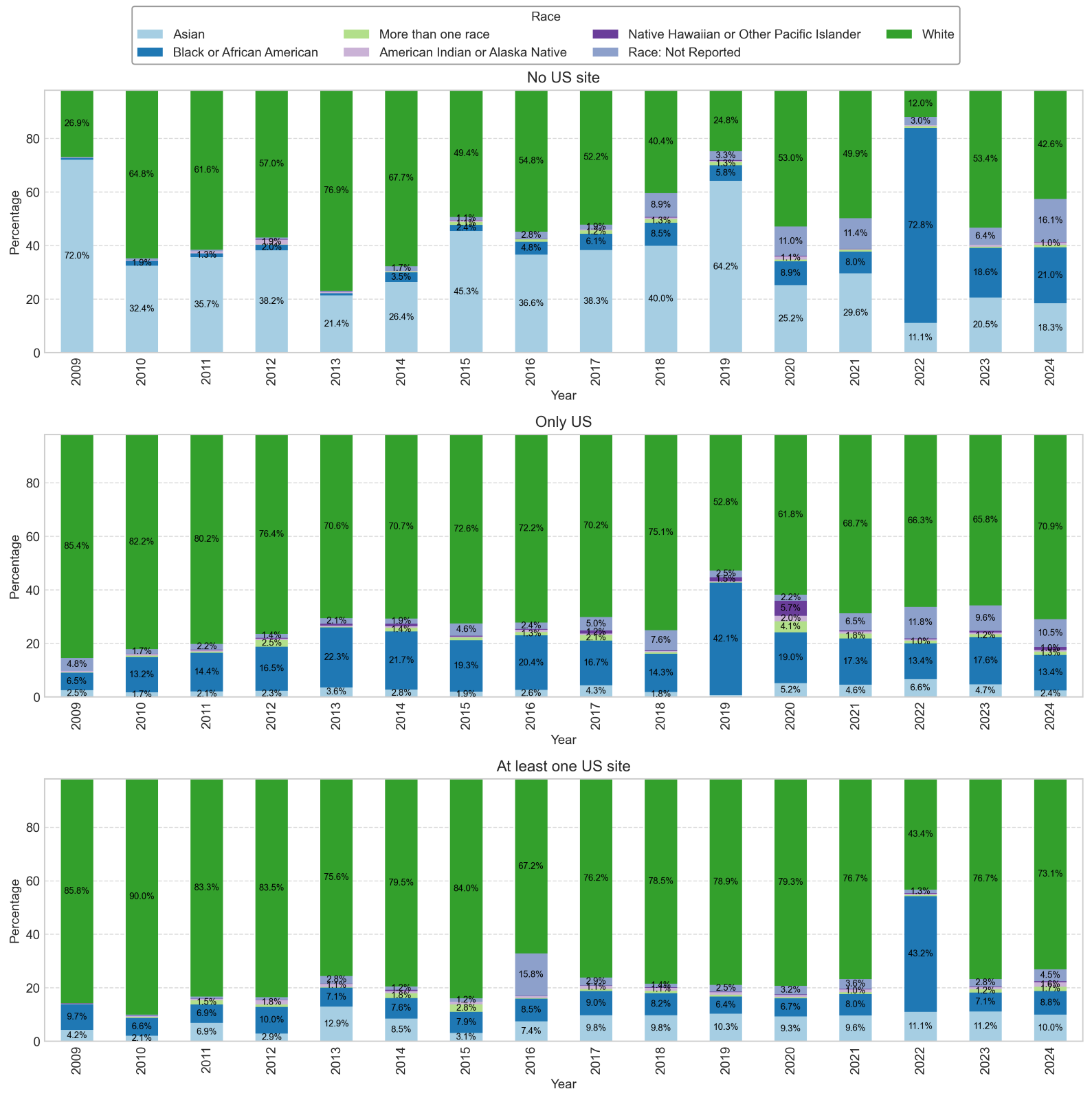
