## Supplementary figures and images for "Racial and Ethnic Diversity in Clinical Studies Reported to ClinicalTrials.gov, 2009-2024"

### Examples of custom race and ethnicity tables from clinicaltrials.gov

NCT03109184

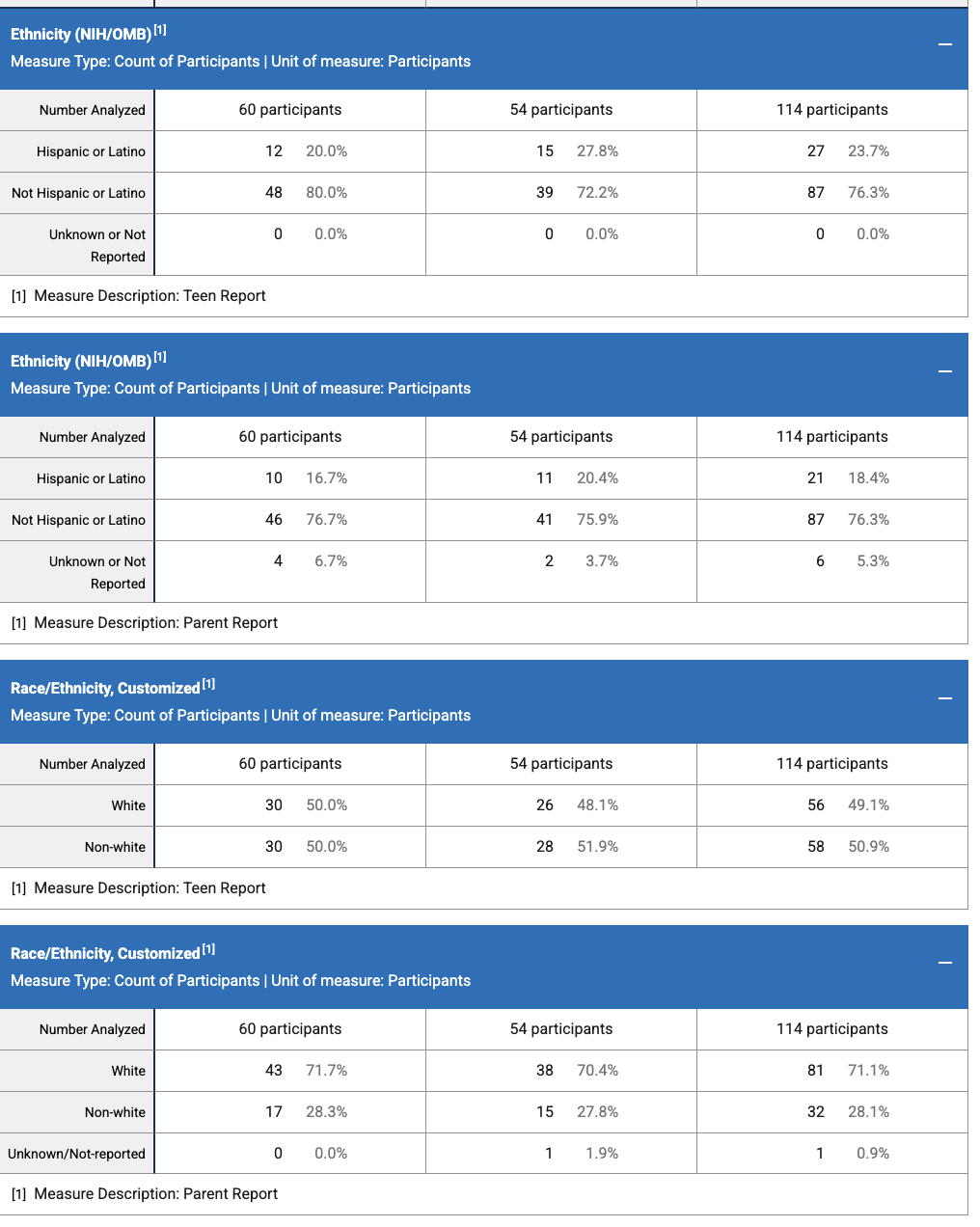

NCT02608684

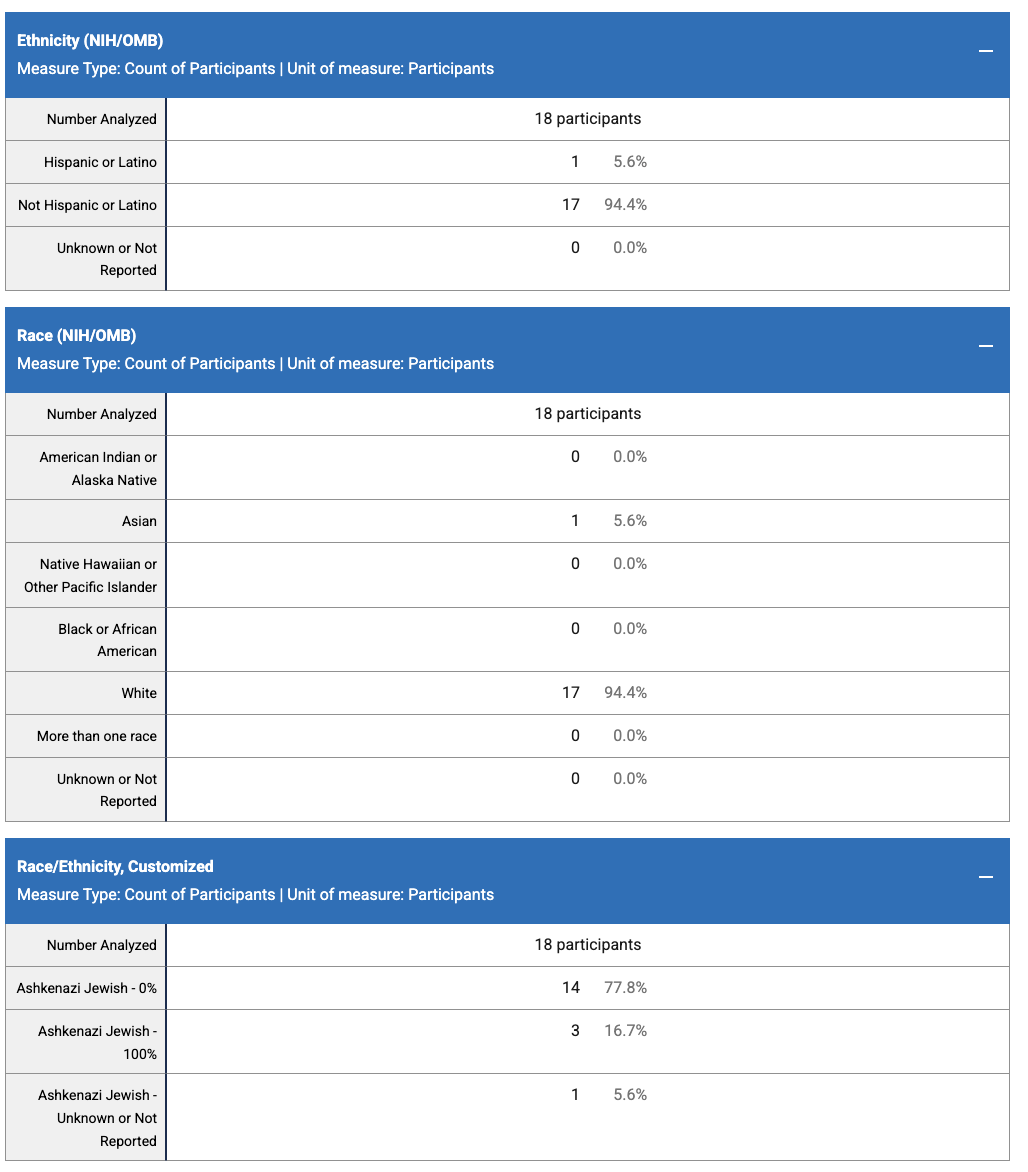

NCT02214121

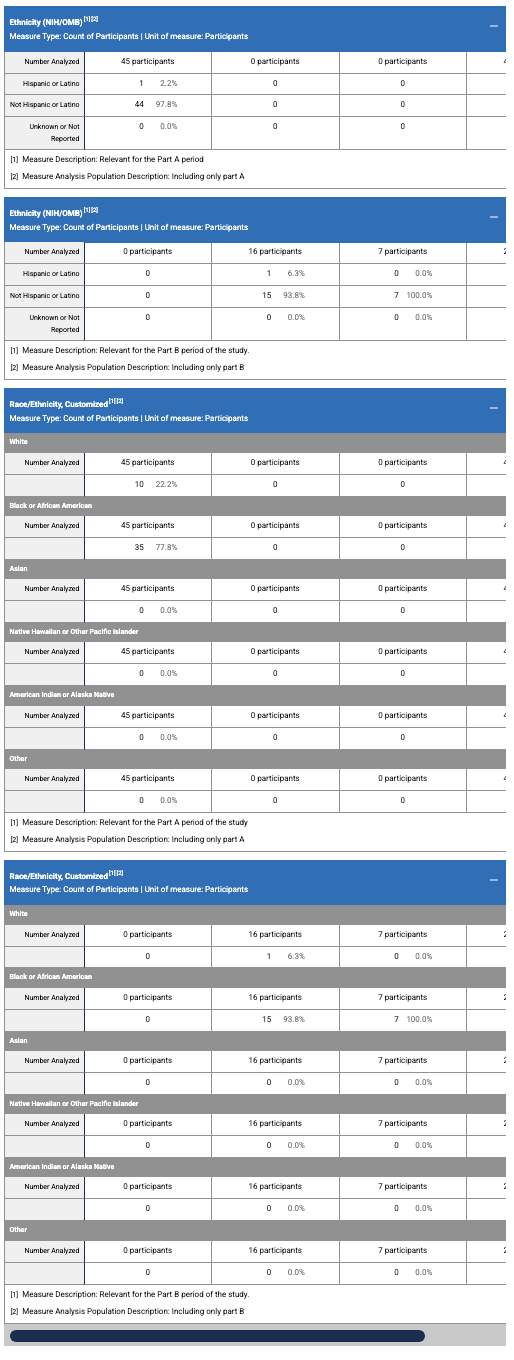

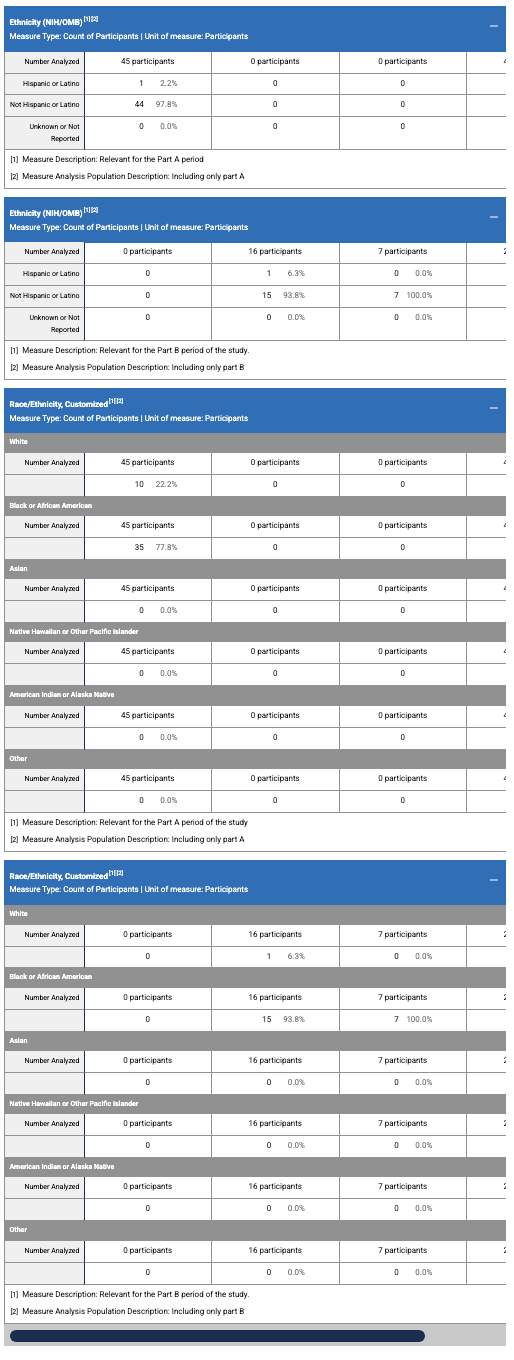
